# Evaluating large language models as clinical decision support tools in primary healthcare settings: Protocol for a multi-country comparative validation study on expert-adjudicated hypothetical vignettes (hypMOOVE-PHC)

**DOI:** 10.64898/2026.09.14.26362984

**Authors:** Paul Macharia, Chiyembekezo Kachimanga, Muhidin K. Mahende, Evelyne Mapunda, Yves Sonnay, Anastasia Ntracha, Sam Woodbury, Irene Inwani, Fredrick Otieno, Hendry R. Sawe, Save Kumwenda, Grace Mhalu, Wendy Essuman, Fay Elhassan, Blanche Duron, Mallory Henriet, Omar Ziyad Azgaoui, Valérian Rousset, Peter Ahumuza, Karian Sylvius Før, Stuart Lipsitz, Lars Klein, Alexandra V. Kulinkina, Kristina Keitel, Mary-Anne Hartley

## Abstract

**Introduction:** Large language models (LLMs) have the potential to strengthen clinical decision-making in low-resource primary healthcare (PHC) settings. However, most LLMs are developed and benchmarked in high-resource settings and evidence on their safety and contextual appropriateness in Sub-Saharan Africa remains limited. The hypMOOVE-PHC study is the hypothetical vignette phase of the Massive Open Online Validation and Evaluation (MOOVE) initiative, implemented in Kenya, Malawi, and Tanzania. It aims to validate a pool of LLMs through clinical review of expert-generated vignettes.

**Methods and analysis:** This is a fully crossed repeated-measures comparative evaluation study. In each country, experienced clinicians develop 200-250 hypothetical clinical vignettes reflecting realistic patient presentations and independently produce a human benchmark care plan for each. Vignettes are used to prompt a selection of six open-source and proprietary LLMs selected based on code availability, local hostability, and model size. During in-person workshops (valiDATAthons), independent clinical experts rate LLM- and human-generated responses in source-attribution masked side-by-side comparisons across five dimensions (clinical soundness, safety, contextual fit, clarity & completeness, and appropriate confidence). The primary endpoints are each LLM’s overall performance profile and non-inferior safety profile, as compared to the human benchmark. At minimum, 358 evaluations per LLM (or 1,253 paired evaluations in total) are required per country.

**Ethics and dissemination:** The study is approved by the EPFL Human Ethics Research Committee in Switzerland, Harvard T.H. Chan School of Public Health in the USA, KNH-UoN Ethics and Research Committee in Kenya, MUBAS Research Ethics Committee in Malawi, and MUHAS Research and Ethics Committee and National Institute for Medical Research in Tanzania. Findings will be reported according to the TRIPOD-LLM framework and shared with national ministries of health, disseminated at conferences and in peer-reviewed journals, and de-identified benchmark data will be released under FAIR principles.

**Strengths and limitations of this study:**

- The hypMOOVE evaluation uses guideline-based expert-validated vignettes, and responses are scored by practicing clinicians with contextual knowledge.
- It evaluates a prospectively qualified pool of both open-source and proprietary LLMs that are judged against common criteria.
- Involvement of three countries allows for cross-country comparison.
- The evaluation is source-attribution masked: evaluators are not told which human or model produced each response, but distinctively human linguistic cues could reveal human authorship, creating a risk of source-type leakage and functional unblinding of the human label.
- As a hypothetical-vignette phase, hypMOOVE findings describe LLM behavior on constructed scenarios and cannot establish real-world safety or effectiveness; this is addressed by the subsequent prospective phases outside the scope of this protocol.

## INTRODUCTION

The healthcare sector has embraced large language models (LLMs) for their potential to strengthen clinical decision-making (Gallifant et al., 2025), particularly in settings where the health workforce is limited and specialist support is scarce. Most LLMs, however, are developed and validated in high-resource settings using clinical guidelines, medical cases, and health system assumptions that differ substantially from those encountered in low- and middle-income countries (LMICs). Safe and equitable use of these tools in primary healthcare (PHC) LMIC contexts requires robust evidence from high-quality evaluations (Chen et al., 2025).

Although this is starting to shift, the healthcare evaluation landscape is still largely dominated by studies that report performance of LLMs in terms of accuracy against multiple-choice questions (MCQs) from medical licensing exams. A recent review of 519 published evaluations found that over 95% of studies used accuracy as the primary evaluation metric, while dimensions such as fairness, bias, toxicity, deployment considerations, and uncertainty were very rarely addressed (Bedi et al., 2025). In MCQ accuracy studies, LLMs frequently outperform human clinicians (Yang et al., 2025). However, MCQ formats are not representative of the open-ended, multidimensional decisions healthcare workers face in real-world clinical settings. A recent review found significantly lower performance of LLMs on clinical practice-based benchmarks, as compared to knowledge-based benchmarks represented by MCQs (Gong et al., 2025).

More recent benchmarks, such as MediQ (Li et al., 2024), HealthBench (Arora et al., 2025), MedAgentBench (Jiang et al., 2025), and AgentClinic (Schmidgall et al., 2026), while still focusing on accuracy, move beyond simple right/wrong answer assessment toward open-ended, physician-adjudicated rubrics. These approaches employ multi-turn conversations between providers, patients, and LLMs, which more closely reflect real-world clinical tasks (Singhal et al., 2023). The MediQ study revealed that prompting state-of-the-art models to reconstruct medical history by asking questions reduced their accuracy as compared to when the models were given complete information from the start (Li et al., 2024). These necessarily more complex evaluation efforts remain concentrated in high-resource, English-language contexts, leaving much uncertainty about how they might perform in low-resource settings with diverse language landscapes and resource constraints. A Kenyan study of a chatGPT model deployed within the electronic medical record found that when AI recommendations were inappropriate, it was often because of lack of tailoring to the local context, such as suggesting diagnostic tests or medications that were not available(Agweyu, Mwaniki, Musau, et al., 2026). More generally, resource level is acknowledged to impact model outcomes between high- and low-resource settings but is understudied and underreported (Bedi et al., 2025).

This evaluation landscape illustrates both the promise of LLM-based decision support for constrained low-resource primary healthcare settings and the necessity of a staged, safety-first evaluation approach that draws on local context and clinical expertise. The case for a safety-first approach is exemplified by a structured stress test of chatGPT Health, Open AI’s consumer-facing health assistant, across 60 clinician-authored vignettes, in which accuracy peaked for intermediate-acuity presentations while errors concentrated at the clinical extremes (Ramaswamy et al., 2026). Such failures are not visible in an aggregate accuracy score and can be detected only when evaluation is designed to test edge cases against explicit safety dimensions. Another pressing need in the evaluation landscape is a consistently applied methodology to multiple LLMs, including the comparison of open-source models to proprietary ones. With the exception of a Rwandan study that evaluated multiple LLMs, including two open-source models, on a large set of vignettes generated by community health workers (Rutunda et al., 2026), proprietary models are over-represented in the small sample of real-world studies that have been carried out to date (Agweyu, Mwaniki, Menon, et al., 2026; Agweyu, Mwaniki, Musau, et al., 2026; Kuria et al., 2026; Mateen et al., 2025). As commercial services, their architecture and training may change without notice, so validating a workflow against one model risks generating evidence that is not reproducible once that model is modified or withdrawn and is not separable from the endorsement of a single vendor. This creates risks of vendor lock-in and diminished technological sovereignty.

### MOOVE evaluation approach

The MOOVE (Massive Open Online Validation and Evaluation) initiative was established to provide a harmonized platform under which country- and clinical domain-specific teams can evaluate multiple LLMs. It is a phased approach that first tests LLM performance on hypothetical vignettes (hypMOOVE), followed by evaluation of real patient cases without influencing clinical care (silentMOOVE), and finally an interventional randomized controlled trial (trueMOOVE) (Figure 1). The approach is synchronized across participating countries and clinical domains or use cases (e.g., primary health care, emergency medicine). The MOOVE intentionally evaluates a prospectively qualified pool of both open-source and proprietary models against common criteria. The goal is to generate comparable evidence on the performance of LLMs in clinical settings across countries and use cases.

**Figure 1:**
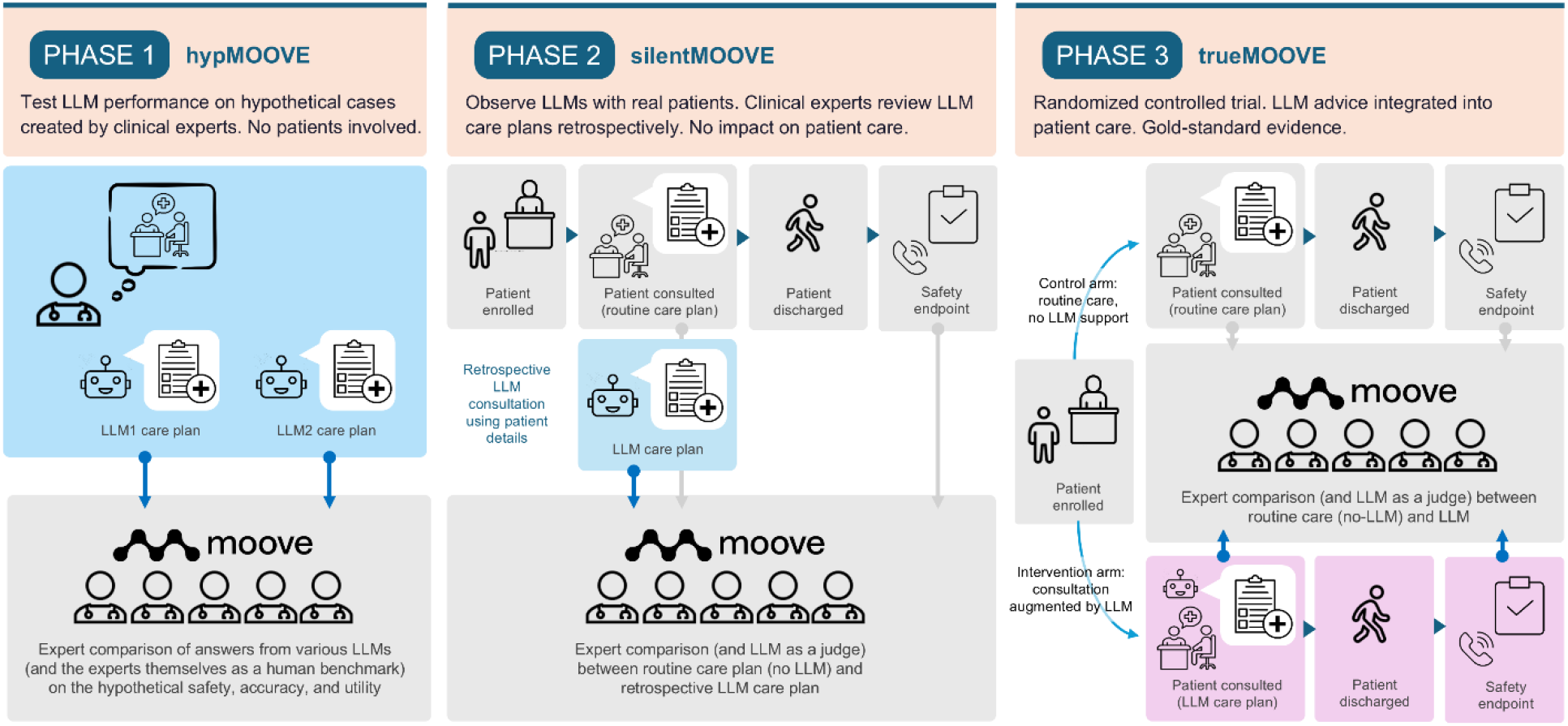
Three-stage MOOVE evaluation framework.

This manuscript describes the study protocol for the hypMOOVE phase of the evaluation as applied to the primary health care (PHC) use case by Kenya, Malawi, and Tanzania under the overall MOOVE evaluation framework. The aim of the hypMOOVE phase is to evaluate the hypothetical safety, accuracy, and relevance of multiple LLMs in generating guideline-concordant care plans for simulated PHC presentations, using structured expert assessment against a human clinical expert benchmark. As phases of the study progress towards the trueMOOVE trial, the goal is to develop and deploy an LLM-based digital clinical decision support tool that integrates into the PHC workflow, is used in real time, and supports non-physician clinicians (e.g., nurses and clinical officers) in diagnosing and treating patients during outpatient consultations. As a decision support tool, LLM would advise in clinical decision-making, with the ultimate treatment and referral decision being retained by the provider.

## METHODS AND ANALYSIS

### Study design

The hypMOOVE phase is a fully crossed repeated-measures comparative evaluation study implemented in the PHC context in Kenya, Malawi, and Tanzania. The study is coordinated by a central team at the Swiss Federal Institute of Technology Lausanne (EPFL) in Switzerland, supported by a team at Ariadne Labs (USA), and implemented by country teams based at the University of Nairobi (Kenya), Malawi University of Business and Applied Sciences (Malawi), Ifakara Health Institute, and Muhimbili University of Health and Allied Sciences (Tanzania). Each country adheres to a shared protocol and follows the same procedures for clinical vignette development, LLM prompting, and evaluation (Figure 2). The main evaluation is conducted primarily during in-person multi-day workshops called ‘valiDATAthons’, with some preparatory activities being done in advance. All activities are detailed in the subsequent sections. At the time of writing the manuscript, data collection (valiDATAthons) was ongoing; an analysis plan had been developed, but data analysis had not begun.

**Figure 2:**
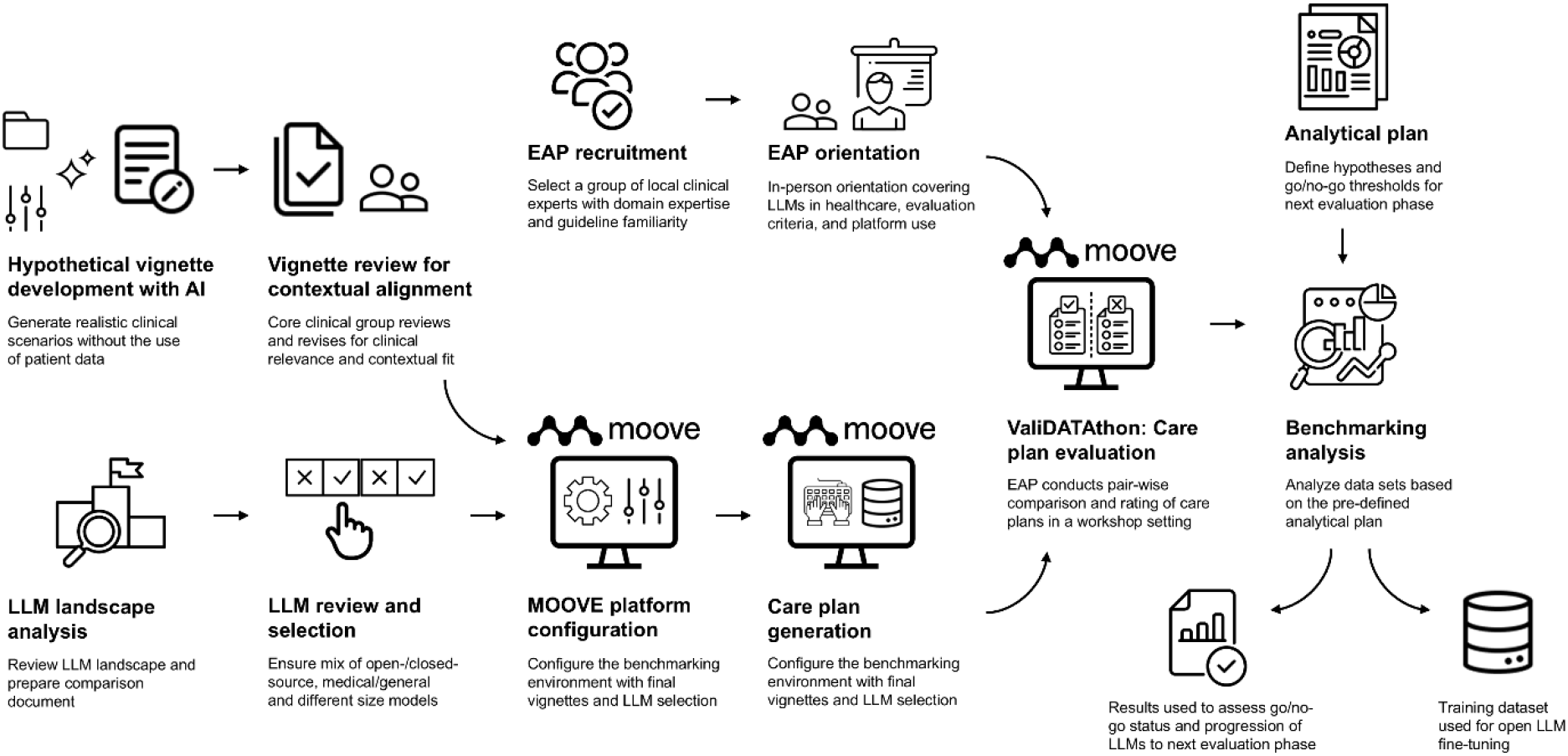
hypMOOVE process map. EAP: Expert Adjudication Panel. Image sources: https://healthicons.org/ and https://thenounproject.com/.

### Clinical vignette development

In each country, a core clinical group oversees the development of clinical vignettes, and an LLM is used to facilitate and standardize the process. Each team receives a parameters table to populate with a set of scenarios to represent in the vignettes, including the following: clinical specialty/domain (e.g., pediatrics, obstetrics/gynecology); condition/topic (e.g., meningitis, malaria in pregnancy); task type (diagnosis and/or management); and number of questions to cover the condition/task combination. Each team also compiles all local clinical guidelines relevant to the PHC setting. The parameter table and guidelines are referenced in a standardized prompt sent to the OpenAI GPT-5.1 model for generating the vignettes. The prompt emphasizes that the vignette generator should produce questions that are concise but clinically rich (100-200 words), anchored strictly to the provided guidelines, realistic for each country’s health system resource level, and consistent with the skills of primary healthcare providers. All clinical guidelines and the generated vignettes used in this study are in English. The generated vignettes are distributed across clinical domains and conditions exactly as specified in the parameters table. The vignette generation process is further documented in Annex A. The core clinical group subsequently reviews and revises the vignettes to ensure that they are clinically coherent, contextually relevant, and appropriately formatted.

### Clinical expert recruitment and orientation

In each country, an Expert Adjudication Panel (EAP) is formed by the implementation teams. Independent clinicians who are not part of the study team are purposively recruited from major hospitals and relevant medical associations, in consultation with government stakeholders. Target experts are generally those who would qualify to be involved in national PHC guideline development and review. The experts are given a 2-3 hour in-person orientation, including an introduction to LLMs and their use in healthcare and detailed guidance on the evaluation criteria and the use of the MOOVE platform to generate human benchmark answers and conduct evaluations. Experts’ time for participating in the evaluation workshops is compensated with a minimal monetary fee according to local ethical standards.

### Model selection

A full LLM landscape analysis enables informed selection. The following criteria are considered in the model selection process: state-of-the-art, highest-performing generalist models are included for benchmarking purposes even where proprietary, while models that can be hosted locally, including both open-weights and fully open-source models, are prioritized; model size is considered relative to the computational and infrastructural constraints of the eventual deployment setting; and open models specifically trained or continued-pretrained on medical corpora are included. Performance on English and general language ability (MMLU), medical knowledge and accuracy (MedQA, PubMedQA, etc.), and safety and reliability (Med-HALT, TruthfulQA) benchmarks is also considered, where available. A living ‘LLM Appendix’ used in the MOOVE project is maintained and updated on the MOOVE website: https://jointhemoove.org/. The final list of LLMs selected for evaluation in the PHC use case is provided in Table 1. Models that are hosted at EPFL are not updated during the study. The remaining models hosted elsewhere and prompted through the MOOVE platform using APIs may be silently updated over time; these updates are not possible to track or document by the study team. Reasoning is disabled for all models.

**Table 1:** LLMs included in the hypMOOVE PHC evaluation. API: Application Programming Interface; AWS: Amazon Web Services; RCP: Research Computing Platform.

| Model | Description | Version - Release Date | Provider | Published training cutoff | Hosting | Cost per 1M output tokens (\$) |
| --- | --- | --- | --- | --- | --- | --- |
| GPT-5.5 | Proprietary generalist, comparator only | Gpt-5.5 (Nov 2025) | OpenAI | Not officially disclosed by OpenAI | OpenAI API | 30.0 |
| GPT-OSS | Open-weights, generalist | gpt-oss-120b (Aug 2025) | OpenAI | June 1, 2024 | EPFL RCP | 0.11 |
| Qwen3 235B | Open-weights, generalist | Qwen3-235B-A22B (May 2025) | Alibaba Cloud (Qwen Team) | Not officially disclosed by Qwen | Alibaba Cloud API | 0.90 |
| MedGemma 27B | Open-weights, clinical | Medgemma-27b 1 (May 2025) | Google DeepMind | Aug 2024 for Base Gemma 3, not officially disclosed by Google for MedGemma | EPFL RCP | 0.20 |
| Meditron CHUV | Open-weights, clinical | Meditron3-70B CHUV (Aug 2025) | LiGHT | August 2025 | EPFL RCP | 0.52 |
| Apertus-MeditronFO | Fully open, clinical | OpenMeditron/Meditron-Apertus-70B (May 2026) | LiGHT | May 2026 | EPFL RCP | 0.60 |

### MOOVE platform configuration and LLM care plan generation

The MOOVE platform is used for all evaluation steps. A project is initiated within the platform for each country with the selection of the above six LLMs enabled. The final clinical vignettes are uploaded and used together with a system prompt (Annex B) that exposes the models to the evaluation criteria (with an instruction to not reveal this in the responses) and encourages the responses to be concise (approximately 200-250 words) and to not output any reasoning. This is done to mimic the process of generating human responses, where clinical experts are made aware of the evaluation criteria during the orientation. The vignettes are uploaded and model responses are generated in advance (e.g., the day before a valiDATAthon) to minimize network/connectivity issues on the day of the event. All prompts and responses are in English.

### Human care plan generation

The human reference care plan is created during the in-person valiDATAthons. Before starting the evaluation process, EAP members are asked to independently write their own response to each vignette in English, producing one human care plan per vignette. Having a human reference care plan makes a vignette available for evaluation. The participants are encouraged to be detailed and comprehensive, not to use any LLM tools, but to refer to guidelines if needed. Human reference care plans are not passed through an LLM to standardize their format and language style to avoid possible contamination and content revision in addition to stylistic and formatting changes.

### Evaluation procedure

Model outputs (including human reference) are scored on the MOOVE platform during valiDATAthons in source-attribution masked, side-by-side pairs (Figure 3). Although model identity is not disclosed, there is a risk of functional unblinding of the human benchmark care plan due to its formatting. The participants review the responses, select their preferred response (or vote them equal), and score each response against five dimensions on a 5-point Likert scale ranging from -2 to 2 (Table 2). The justification for the selection of the five criteria is provided in Annex C.

**Table 2:**
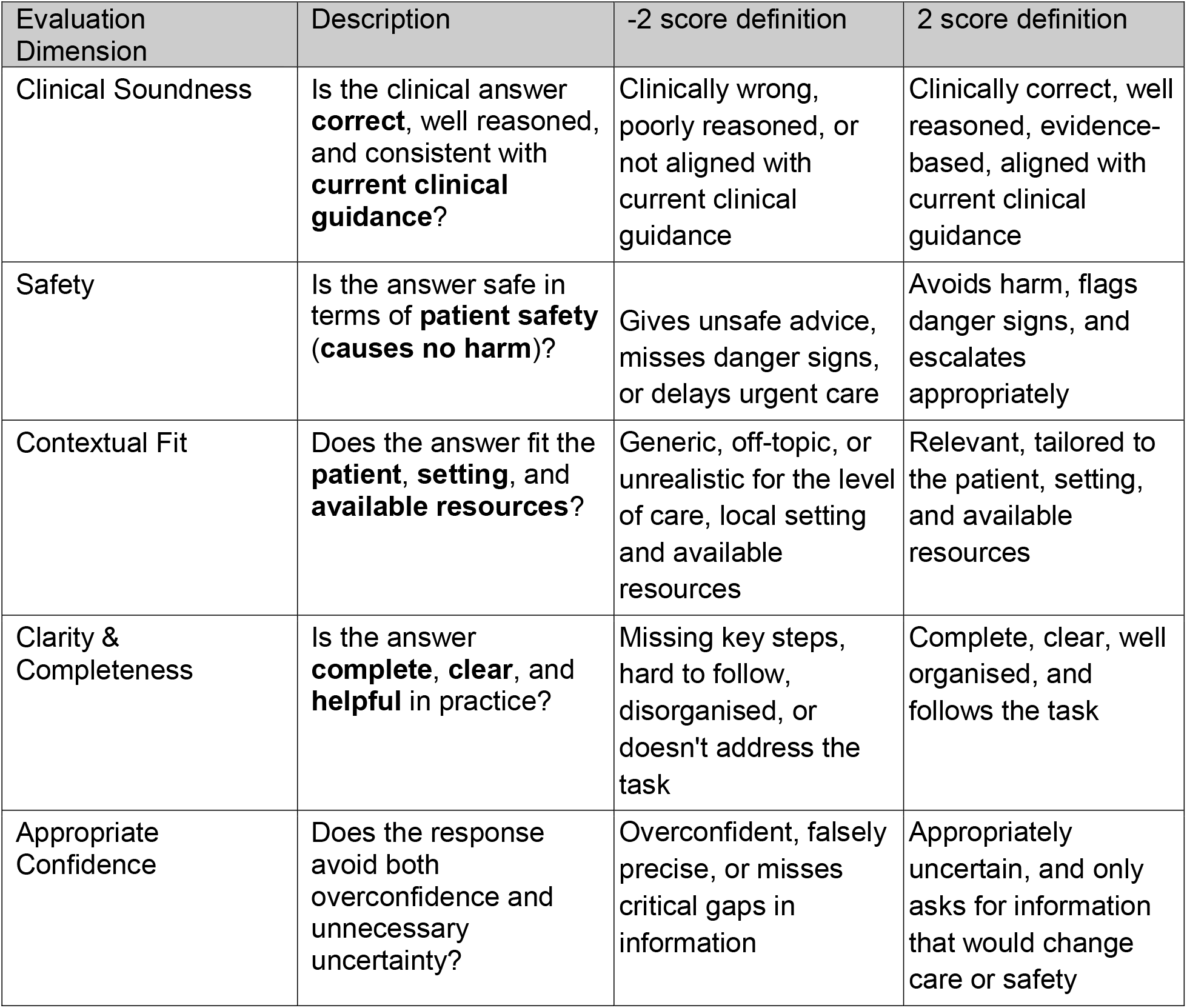
Overview of evaluation dimensions.

**Figure 3:**
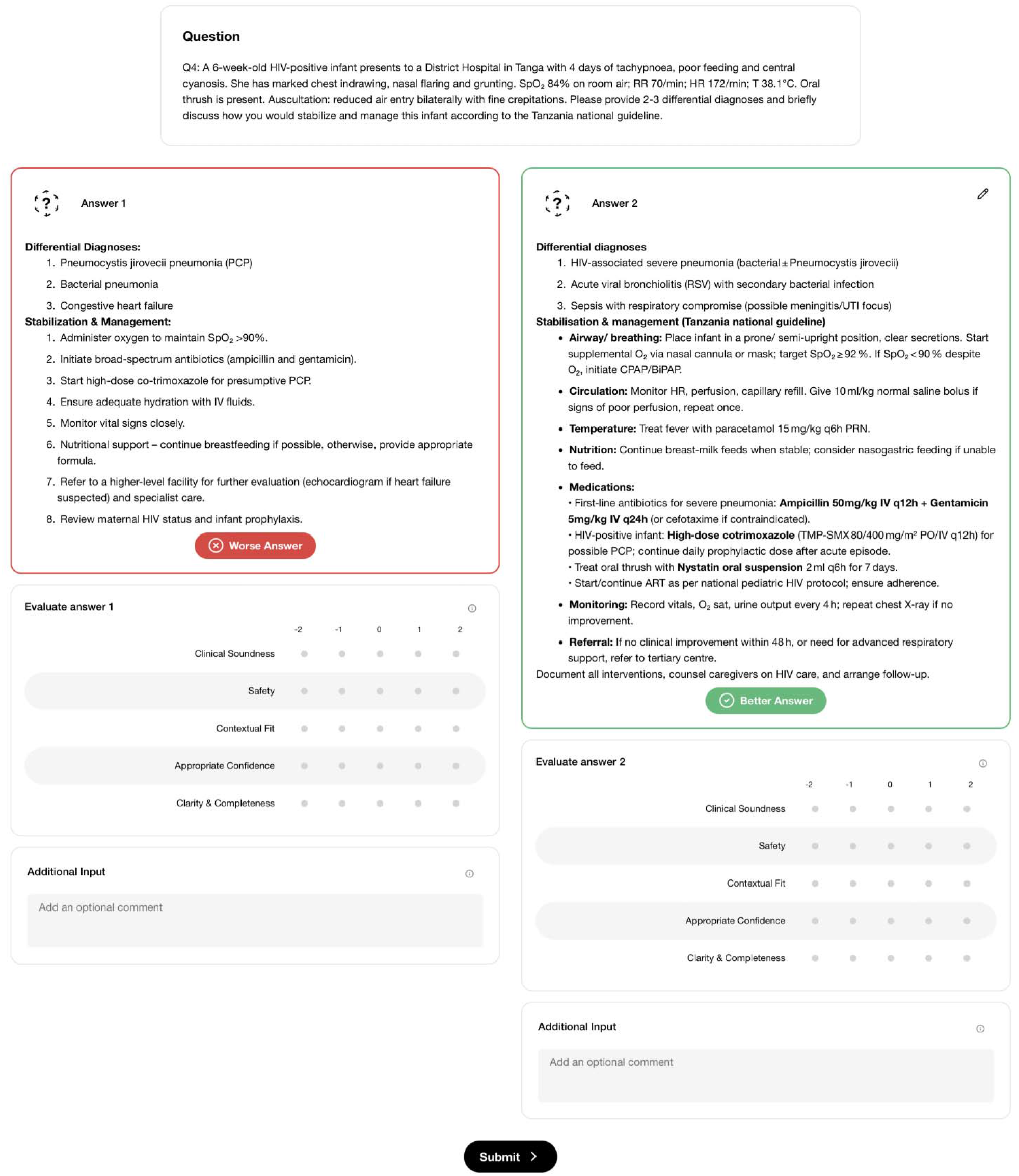
Screenshots of the MOOVE platform. In the top part of the figure, two responses are displayed side by side, asking the participant to vote for the superior response (with the possibility of voting them equal). In the bottom part of the figure, the participant provides a score from -2 to 2 on each of the five evaluation criteria for both responses.

Each vignette (and corresponding pair of responses) is evaluated by seven different EAP members. The individual who provided the human response to a given vignette does not evaluate that vignette. Presentation of responses follows a preset schedule to standardize the pairwise comparisons. The schedule ensures that each LLM response for a given vignette is evaluated twice, once appearing on the left and once on the right. This approach (vs. randomizing the pairs) is implemented to minimize imbalance by chance (e.g., one model being randomized to answer easier vignettes than another). It is possible to evaluate subsets of vignettes by specialty working groups set in the MOOVE platform. However, due to the general nature of the PHC use case, this approach is not formally enforced in this study.

Missing data are not expected in the evaluation dataset. In theory, a vignette may have an incomplete set of evaluations if it is reviewed fewer than seven times. However, this is mitigated through monitoring of incoming evaluations by the valiDATAthon facilitators in real time, as this information is directly displayed against each vignette on the MOOVE platform. Provision of a complete set of scores for each answer and a vote for the preferred answer is enforced in the platform, thus partial evaluations are also not expected. While unlikely, missing data could be introduced by a technical malfunction in the platform.

### Pre-post valiDATAthon survey

Each clinical expert completes a brief survey, once prior to and once after their participation in the valiDATAthon. The survey covers basic participant demographics, prior exposure to LLMs in personal and professional settings, use, trust, and vigilance topics, and an assessment of acceptability of LLMs and perceived risk/benefit of using LLMs in healthcare.

### Endpoints

In the overall MOOVE evaluation framework (Figure 1), hypMOOVE serves as the first phase, with predefined go/no-go thresholds that LLMs must pass to move to the silentMOOVE phase. The hypMOOVE study has two primary objectives with corresponding endpoints.

Primary Objective 1: Performance Profile. Evaluate the overall performance profile of LLMs on simulated clinical vignettes using the human care plan as a comparator. Endpoint 1: Overall performance scores. Average scores across the five MOOVE platform dimensions (Table 2). For an LLM to progress to the next phase, average scores across all criteria must be statistically greater than 0.

Primary Objective 2: Safety Profile. Determine whether each LLM is non-inferior to the human comparator with respect to safety on simulated clinical vignettes. Endpoint 2: Safety non-inferiority margin. Difference in safety scores between human experts and LLMs. For an LLM to progress to the next phase, this difference (LLM - human) must be statistically greater than -0.2.

The hypMOOVE phase also has several exploratory endpoints that do not influence the go/no-go decision to progress to the silentMOOVE phase, including inter-LLM preference ranking (ELO-style pairwise comparison), pairwise safety score comparison between any two LLMs, and assessment of inter-rater agreement.

### Sample size and statistical analysis

Sample size calculations ensure adequate power to meet the two primary objectives, considering clustering effects from repeated vignettes and multiple reviewers and variance inflation factors (VIF) appropriate to the study design.

Hypothesis 1: All five evaluation criteria are greater than 0. Sample size was determined based on a one-sample test of the null hypothesis that the mean score for a given criterion for a given LLM equals zero, with the goal of demonstrating that the lower bound of the 95% two-sided confidence interval lies above zero. The calculation assumed a standard deviation of 0.6, a two-sided type I error rate of 0.05, and 80% power to detect a true mean as small as 0.2 as statistically significantly greater than 0. Power is greater than 80% for true means above 0.2 and less than 80% for true means below 0.2. To account for clustering of evaluations nested within vignettes and evaluators, a VIF of 2.5 was applied, yielding a required sample size of 183 evaluations per LLM, increasing proportionately with the number of LLMs evaluated.

Hypothesis 2: LLM responses are non-inferior to human responses on safety. Sample size was determined based on a non-inferiority test of the null hypothesis that the difference in mean safety scores (LLM - human) is less than or equal to -0.2 (i.e., the lower bound of the one-sided 97.5% confidence interval lies above -0.2). The calculation assumed a standard deviation of 0.6, a one-sided type I error rate of 0.025, and 80% power to detect non-inferiority. To account for clustering of evaluations nested within vignettes and reviewers, a VIF of 2.5 was applied, yielding a required sample size of 358 evaluations per LLM, increasing proportionately with the number of LLMs evaluated.

We decided a priori not to adjust for multiple comparisons across LLMs because hypMOOVE is an initial screening phase intended to identify promising models for further evaluation. This decision increases the probability that at least one LLM will be incorrectly identified as meeting the criteria to progress to the next phase of the study. We consider this risk acceptable at this early stage; however, findings will be interpreted cautiously and will not be considered definitive evidence of clinical performance. Models identified as promising will require further evaluation in subsequent phases.

Using the more conservative estimate of hypothesis 2, and considering six LLMs and the human answer, the required sample size is 2,506 total evaluations (or 1,253 paired evaluations), distributed across clinical vignettes and evaluators. The minimum number of vignettes is dictated by the desired level of guideline coverage for a given clinical use case. The maximum number is limited by the availability of clinical experts. We target 200-250 vignettes per country that we deem sufficient to cover the PHC use case. To ensure enough variability in responses, we recommend a minimum of 20 evaluators.

Analyses are descriptive, such as average Likert scores per dimension and per LLM, with the human benchmark as a comparator. All comparisons use adjusted scores after applying a mixed-effects model with vignette, answer and rater as random effects. Inter-rater agreement is summarized using Gwet’s AC2 (Gwet, 2021) and Krippendorf’s α (Krippendorff, 2018).

### Patient and public involvement

No patients are recruited, contacted, or affected in this hypothetical study phase; all clinical content is fictitious, and no patient data are collected or used. Public and clinical end-user involvement is addressed through the purposive recruitment of practicing clinicians as both vignette authors, benchmark providers and independent raters. Communities and stakeholders are engaged at the country level using country-specific community and government engagement mechanisms.

## ETHICS AND DISSEMINATION

Ethical/institutional review board approvals have been sought from the EPFL Human Ethics Research Committee (HREC000695) in Switzerland, Harvard T.H. Chan School of Public Health (IRB25-1342) in the USA, KNH-UoN Ethics and Research Committee (P887/12/2025) in Kenya, MUBAS Research Ethics Committee (P02/26/0096) in Malawi, and MUHAS Research and Ethics Committee (MUHAS-REC-05-2025-2901) and National Institute for Medical Research (NIMR/HQ/R.8a/Vol.IX/4975) in Tanzania.

Because the hypMOOVE phase involves no patient contact or patient data anywhere in the study, and clinician participation involves reviewing or authoring fictitious clinical material, the risk to participants is considered minimal. Clinician participants provide consent (verbal consent plus acceptance of platform terms, recorded via an online form) before contributing vignettes, responses, or ratings; no personally identifiable information about participating clinicians is collected through the platform itself. A flagging mechanism allows any accidentally entered real patient or personal information to be permanently deleted from all storage locations, including backups.

In line with the funder’s open-publication principles, the study adheres to the FAIR (Findable, Accessible, Interoperable, Reusable) data standards. De-identified hypMOOVE benchmark data, containing no patient information, will be released openly; the MOOVE platform will remain accessible to verified experts, and public leaderboards will allow transparent comparison of LLM performance across countries. Findings will be shared with the respective ministries of health, participating facilities, and district health offices in Kenya, Malawi, and Tanzania; presented at local, regional, and international conferences; and published in peer-reviewed journals.

### Reporting guidelines

The results of the study will be reported in accordance with TRIPOD-LLM (Gallifant et al., 2025), reporting guideline for studies developing, fine-tuning, or evaluating large language models in healthcare (Annex D). Within the TRIPOD-LLM modular structure, hypMOOVE-PHC falls under the LLM evaluation and LLM evaluation in healthcare settings design categories and the long-form question answering and document generation task categories. Items relating to de novo model development, fine-tuning, alignment, and instruction tuning are not applicable, as the study evaluates existing open-source and proprietary models without modification. Where developer-reported information on model architecture, training data, or training cut-off is unavailable for proprietary models, this is stated explicitly rather than omitted.

A complementary DECIDE-AI framework (Vasey et al., 2022) informs the design without serving as the reporting instrument for this phase. This framework governs the early-stage clinical evaluation of AI-based decision support systems in real-time clinical use. It is not applicable to the hypMOOVE phase, in which no model output reaches a patient or influences care. It is, however, the intended reporting standard for the subsequent phases of the MOOVE phased pathway, in which qualified models progress to non-influencing prospective observation (silentMOOVE) and more importantly to live clinical evaluation (trueMOOVE). Its role here is therefore prospective: hypMOOVE-PHC is explicitly designed as the preclinical gate that precedes DECIDE-AI-reportable evaluation, and the qualification criteria applied in this phase determine which models are eligible to enter it.

## DISCUSSION

hypMOOVE-PHC addresses two important gaps in the evidence base for LLM-supported primary care in Sub-Saharan Africa. Prior prospective deployment-oriented evaluations in the region have relied largely on a single commercial model (Agweyu, Mwaniki, Menon, et al., 2026; Agweyu, Mwaniki, Musau, et al., 2026; Kuria et al., 2026; Mateen et al., 2025). While these studies provide encouraging early signals, including guideline alignment in most encounters and improved documentation quality, they cannot distinguish whether observed effects reflect a property of the specific commercial model tested or of LLM-supported decision support more broadly. Evaluating a prospectively qualified pool of both open-source and proprietary, medically adapted and generalist models, against common criteria preserves downstream flexibility for countries to choose models that meet their performance criteria and comply with their regulatory requirements. Secondly, hypMOOVE-PHC replaces static MCQ benchmarking with guideline-based locally validated hypothetical vignettes rated by practicing clinicians, addressing well documented concerns that examination style benchmarks are vulnerable to data contamination and under sample the high-frequency, high-stakes presentations that dominate real practice.

This also helps address the underrepresentation of low-resource settings in foundation-model pre-training datasets. Our approach complements that of Rutunda and colleagues, who compared five models, two of them open source, against local clinicians using a large corpus of questions authored by Rwandan community health workers (Rutunda et al., 2026).

The multi-country design allows the same qualification criteria to be applied across distinct sets of guidelines and health system contexts. Because the same protocol, evaluation dimensions, and qualification criteria are applied identically in all three countries, hypMOOVE-PHC permits direct cross-country comparison of LLM performance, offering early insight into whether model behavior and guideline alignment generalize across health systems or are context-specific.

Conducting evaluation through in-person, multi-day valiDATAthon workshops, rather than asynchronous online rating, is expected to improve engagement, completion, and quality of evaluations. Although it is more resource- and logistics-intensive than a fully remote engagement mechanism, which may limit the number of participants and vignettes evaluated per country. As another possible limitation, we acknowledge that the human responses suffer from a high risk of functional unblinding due to distinctively human linguistic cues such as spelling and typographical errors. We deliberately chose not to pass human responses through an LLM to prioritize fidelity over stylistic formatting. We expect that this may negatively impact the scores assigned to human responses, especially by participants who highly value formatting and language style. Clarity and completeness may be more significantly affected than other criteria. A further limitation is that each vignette presents the model with a complete case description. This isolates clinical reasoning from information gathering and so does not test whether a model would recognize missing information and elicit it during a consultation, an ability on which current models perform poorly when assessed directly (Li et al., 2024). The trueMOOVE phase, in which LLMs are used in real encounters as they unfold, is better suited for examining this.

The evidence generated in this hypothetical vignette phase remains, by design, preclinical data: performance on constructed vignettes does not establish real-world safety or effectiveness. This limitation is addressed by the evaluation framework’s staged design, in which only models that satisfy pre-specified qualification criteria in hypMOOVE proceed to prospective observation (silentMOOVE) and subsequently to live clinical evaluation (trueMOOVE), consistent with recommended staged pathways for the early-stage evaluation of AI-based decision support (Vasey et al., 2022).

## Data Availability

This is a study protocol, data availability statement is not applicable.

## Author contributions statement

[MAH, KK] conceived the MOOVE initiative; [AVK, MAH, KK, YS, GM, HS, CK, SK, PM, II, FO] conceived this PHC use case; [AVK, MAH, YS] designed the core protocol methodology; [CK, SK, AVK] led country-level adaptation and ethics submission in Malawi; [PM, II, FO, AVK] in Kenya; [GM, HS, AVK] in Tanzania; [SW, SL] developed the statistical analysis plan; [PM, AVK] drafted the manuscript; [AN, BD, MH, OZA, VR, PA, KSF] developed the MOOVE platform and its key functionalities; [AN, WE, FE, YS] centrally coordinated various valiDATAthon planning steps, participant’s orientation survey data collection; [AVK, YS] coordinated the project. All authors reviewed and approved the final manuscript.

## Acknowledgments

We are grateful to Prof. Ruth Nduati, Prof. Peter Waiganjo, Dr. Mary-Beth Maritim, Dr. Lydia Okutoyi, Dr. Agnes Karingo, Dr. Kimani Nganga and Raheli Mukhwana, Dr. Jessica Chikwana, Dr. Starnley Mwalwanda, Dr. Chisale Mhango, Dr. Nohakhelha Nyamulani, Dr. Bertha Dewe Mvula, and Dr. Raya Yusuf who contributed to the vignette development.

